# Neutralization of SARS-CoV-2 variants by rVSV-ΔG-spike-elicited human sera

**DOI:** 10.1101/2021.11.22.21266673

**Authors:** Yfat Yahalom-Ronen, Noam Erez, Morly Fisher, Hadas Tamir, Boaz Politi, Hagit Achdout, Sharon Melamed, Itai Glinert, Shay Weiss, Inbar Cohen-Gihon, Ofir Israeli, Marina Izak, Michal Mandelboim, Yoseph Caraco, Noa Madar-Balakirski, Adva Mechaly, Eilat Shinar, Ran Zichel, Daniel Cohen, Adi Beth-Din, Anat Zvi, Hadar Marcus, Tomer Israely, Nir Paran

## Abstract

The emergence of rapidly spreading variants of severe acute respiratory syndrome coronavirus 2 (SARS-CoV-2) poses a major challenge to the ability of vaccines and therapeutic antibodies to provide immunity. These variants contain mutations at specific amino acids that might impede vaccine efficacy. BriLife® (rVSV-ΔG-spike) is a newly developed SARS-CoV-2 vaccine candidate currently in Phase II clinical trials. It is based on a replication competent vesicular stomatitis virus (VSV) platform. rVSV-ΔG-spike contains several spontaneously-acquired spike mutations that correspond to SARS-CoV-2 variants’ mutations. We show that human sera from BriLife® vaccinees preserve comparable neutralization titers towards alpha, gamma and delta variants, and show less than 3-fold reduction in neutralization capacity of beta and omicron compared to the original virus. Taken together, we show that human sera from BriLife® vaccinees overall maintain neutralizing antibody response against all tested variants. We suggest that BriLife® acquired mutations may prove advantageous against future SARS-CoV-2 VOCs.

## Introduction

BriLife® is a recombinant replication-competent Severe Acute Respiratory Syndrome Coronavirus-2 (SARS-CoV-2) vaccine candidate, based on Vesicular Stomatitis Virus (VSV) platform, in which VSV glycoprotein (G) was replaced by the SARS-CoV-2 spike protein (S), creating rVSV-ΔG-spike [1].

Since the emergence of the original SARS-CoV-2, several variants have been described, including alpha (B.1.1.7), beta (B.1.351), gamma (P.1), delta (B.1.617.2), and most recently omicron variant (B.1.1.529) (https://www.cdc.gov/coronavirus/2019-ncov/variants/variant-info.html(accessed on January 24th, 2022; Figure 1a)). The emergence of SARS-CoV-2 variants presents a challenge to the ability of vaccines to provide proper immune response.

**Figure 1:**
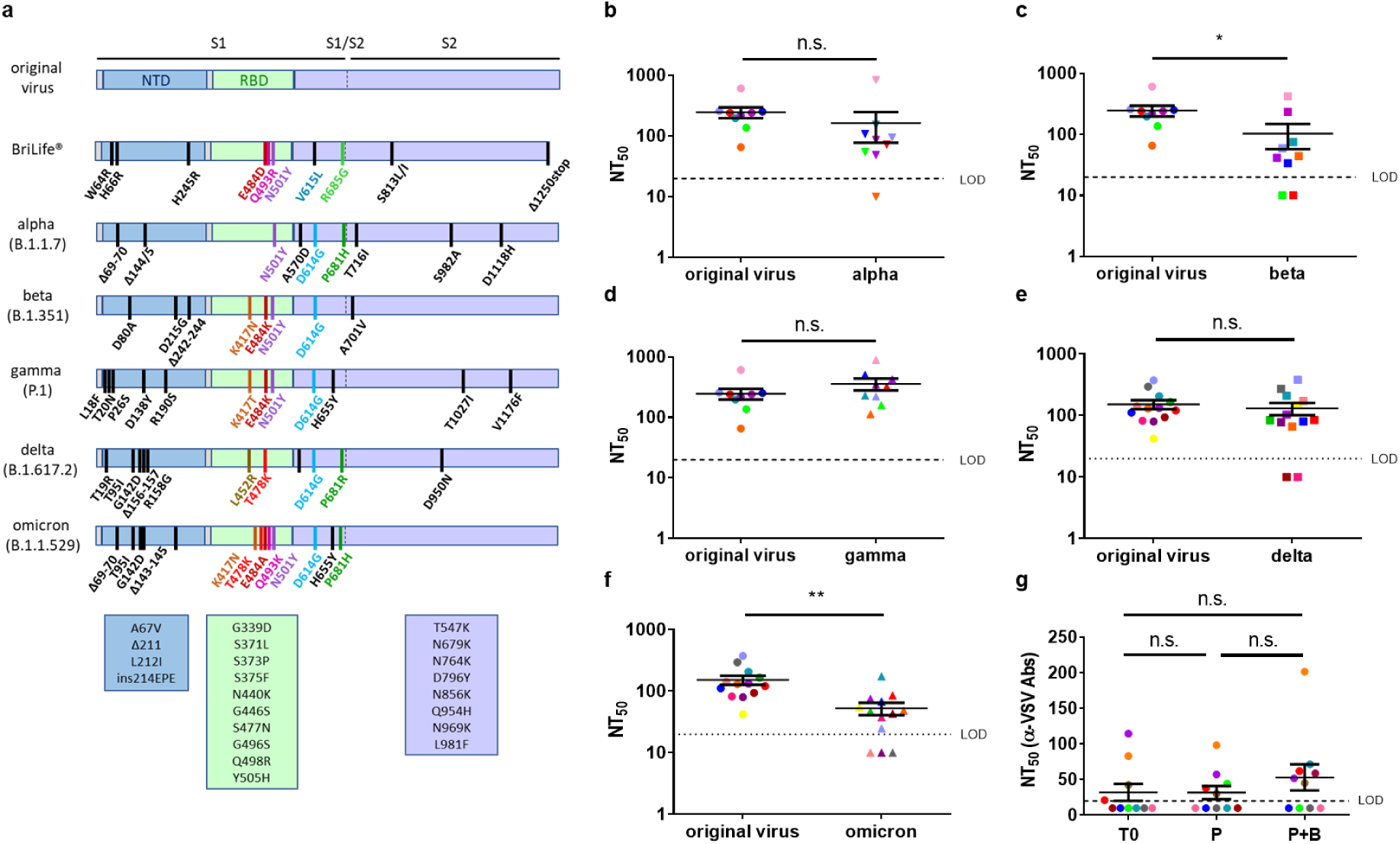
Neutralization of alpha, beta, gamma, delta and omicron variants by sera from BriLife®-vaccinated human participants: (a) Schematic drawing showing spike mutations in BriLife® and in SARS-CoV-2 variants. Main spike domains are indicated. Additional mutations in omicron VOC are listed in rectangles below omicrons’ scheme. Neutralization titers (NT_50_) of BriLife® participants’ sera (following boost vaccination) against SARS-CoV-2 original virus, compared to (b) alpha, (c) beta, (d) gamma (n=9), (e) delta and (f) omicron (n=13) variants. Each serum is color coded for all tested viruses. LOD-limit of detection (LOD=20). Samples below LOD were assigned a value of 10 (half of the LOD). Significance was determined by unpaired t-test. p values: 0.4088, 0.0498, 0.2528, 0.5936, 0.0018 for comparison of original virus to alpha, beta, gamma, delta and omicron, respectively. (g) NT_50_ against rVSV-WT in sera samples of 10 vaccinees collected at time zero (T0), 28 days post 1^st^ vaccination (P) and two weeks following 2^nd^ vaccination (P+B). Sera samples are color coded. Statistical significance was determined by one-way ANOVA nonparametric test with Kruskal–Wallis test. Data is presented as mean ± SEM.

We report here that during its development, BriLife® spontaneously acquired several mutations at specific sites, including but not exclusive to the receptor binding domain (RBD), some of which are identical, or correspond to some of the key mutations or areas of SARS-CoV-2 VOCs, namely N501, E484, Q493 and G685. Here, we examined the neutralization capacity of serum samples from participants in BriLife® phase II clinical trial against the original SARS-CoV-2 virus as well as the alpha, beta, gamma, delta and omicron variants. We show that BriLife®-elicited sera maintain neutralization against SARS-CoV-2 variants. This preservation of neutralization capacity among variant may be attributed to the unique genetic features of BriLife® vaccine.

## Materials and methods

### Samples

Serum samples of BriLife® vaccinees were obtained from participants in a randomized, multi-center, placebo-controlled, dose-escalation phase II of an ongoing clinical trial, aimed to evaluate the safety, immunogenicity and potential efficacy of BriLife®, an rVSV-SARS-CoV-2-S vaccine (IIBR-100) in adults (ClinicalTrials.gov -NCT04608305). All participants were vaccinated intramuscularly in a prime-boost regimen (28 days apart). Samples were obtained approximately two weeks following boost vaccination. This study was performed in compliance with International Council for Harmonization (ICH) Good Clinical Practices (GCP), including the archiving of essential documents as well as the ethical principles of the Declaration of Helsinki.

Serum samples of convalescent COVID-19 patients diagnosed at the first wave of the pandemic were collected by the National Blood Services of “Magen David Adom” in Israel within a protocol for plasma donation. All convalescent volunteers gave their informed consent to the National Blood services of Magen David Adom. The study was approved by the ethics committee of the Israeli Ministry of Health (0083-20-WOMC) [2].

### Cells

Vero E6 cells (ATCC CRL-1586(tm)) were grown in DMEM containing 10% fetal bovine serum (FBS), MEM nonessential amino acids (NEAA), 2 mM L-glutamine, 100 Units/ml penicillin, 0.1 mg/ml streptomycin, 12.5 Units/ml nystatin (P/S/N). Calu3 cells (ATCC HTB-55) were grown in RPMI supplemented with 10% FBS, NEAA, 2 mM L-glutamine, P/S/N, and 1% Na-pyruvate, all reagents are from Biological Industries, Beit-Haemek, Israel. Cells were cultured at 37 °C, 5% CO2 with 95% humidity.

### Viruses

Virus stocks of SARS-CoV-2 original virus (GISAID accession EPI_ISL_406862) were propagated (four passages) in Vero E6 cells. SARS-CoV-2 variants were provided by the Central Virology Lab of the Israel Ministry of health [3]. Alpha (B.1.1.7, GISAID accession EPI_ISL_4169857) was passaged once in Vero E6, followed by two passages in Calu3 cells. Beta (B.1.351, GISAID accession EPI_ISL_4169885), gamma (P.1, GISAID accession EPI_ISL_4169886) and delta (B.1.617.2, GISAID accession EPI_ISL_4169986) variants were propagated in Vero E6 cells. Omicron (B.1.1.529) was propagated in Calu3 cells. Variants were verified by next-generation sequencing (NGS). Recombinant VSV Indiana serotype (rVSV-WT) was propagated in Vero cells [1]. All virus stocks were tittered on Vero E6 cells as previously described [1]. Handling and working with SARS-CoV-2 virus were conducted in a BSL3 facility in accordance with the biosafety guidelines of the Israel Institute for Biological Research (IIBR).

### Plaque reduction neutralization test (PRNT_50_)

PRNT_50_ was previously described [1]. Briefly, serial serum samples were incubated with 300 pfu/ml of SARS-CoV-2 original virus, its variants, or with rVSV-WT. Vero E6 cells were infected with the virus-serum mixtures and following incubation (24 h for rVSV-WT, 72 h for SARS-CoV-2 original virus, beta, gamma or delta variant, 96 h for alpha variant, or 120 h for omicron variant) plaques were counted and NT_50_ was calculated using GraphPad Prism 6 software (GraphPad Software Inc.).

### Statistical analysis

Data were analyzed with GraphPad Prism 6 software. Exact p values are provided for each analysis. Statistical significance for SARS-CoV-2 variants neutralization were determined by unpaired t-test. Statistical significance for VSV vector immunity was determined by one-way ANOVA non-parametric test by Kruskal–Wallis test.

## Results

BriLife® development was accompanied by spontaneous acquisition of several mutations, some of which are at the spike protein at sites of major importance to antibody-mediated immunity, such as E484, Q493, and N501 located in the receptor binding domain (RBD) (Figure 1a) [1]. An additional mutation is R685G at the RRAR multibasic furin cleavage site, located at the junction of receptor-binding (S1) and fusion (S2) domains of the spike protein [1].

We aimed at assessing the neutralization capacity induced by BriLife®. Therefore, we first tested blinded serum samples from two cohorts of participants in a BriLife® phase II clinical trial two weeks following intramuscular prime-boost vaccination for their ability to neutralize SARS-CoV-2 variants alpha, beta, gamma, delta or omicron. Inclusion criteria were negative SARS-CoV-2 nucleocapsid binding (data not shown) and positive SARS-CoV-2 neutralization response against the original virus.

Nine tested sera (“cohort 1”, Table S1) efficiently neutralized the original virus, as well as gamma variant, with mean titers of 246 and 357, respectively (0.7-fold change, Figure 1d, Table S1). Nearly all tested sera neutralized alpha (8/9 samples) and beta (7/9 samples) variants, with mean titers of 163 and 103, respectively (1.5-fold and 2.4-fold reduction relative to original virus, Figure 1b-c, Table S1).

Several months ago, delta variant of concern (VOC) became dominant worldwide exhibiting increased transmissibility, immune escape [4], and breakthrough infections in vaccinated individuals [5]. We therefore tested another set of 13 vaccinees’ sera (“cohort 2”, Table S2) for neutralization of original virus and delta VOC. Of 13 tested sera which neutralized the original virus, eleven also neutralized the delta variant, with no significant difference in neutralization titers (mean titers of 152 and 131, respectively, Figure 1e, Table S2).

Most recently, at the end of November 2021, omicron variant emerged leading to massive waves of infections worldwide, with increasing evidence of breakthrough infections of vaccinated individuals or previously infected individuals [6]. Thus, we also tested the ability of BriLife® vaccinees’ sera (“cohort 2”) to neutralize live omicron VOC. We show that most tested sera (10/13 samples) maintained neutralization capacity against omicron VOC, showing a ∼2.9-fold reduction in neutralization titers (Figure 1f, Table S2), with only 3 vaccinees’ sera below limit of detection (LOD).

Immunity to surface antigens of viral vectored-based vaccines (e.g. adenovirus-based platforms) might hamper their efficacy upon repeated usage. Replacement of VSV-G surface antigen with the antigen of interest reduces the risk of vector immunity [7]. We show that vector immunity was not significantly induced following BriLife® vaccination (Figure 1g, Table S3), minimizing the risk of reduced vaccine efficacy upon repeated vaccinations.

## Discussion

SARS-CoV-2 emerging variants pose a challenge to the battle against COVID-19. Amino acid substitutions in SARS-CoV-2 variants such as delta and omicron were shown to compromise the protection afforded by vaccines, therapeutic antibodies or antibodies derived from previous SARS-CoV-2 infection [8-10]. As described, BriLife® development was accompanied by spontaneous acquisition of mutations, such as E484, Q493, N501 at the RBD and R685G at the furin cleavage site (Figure 1a) [1]. These mutations are similar or identical to some of the key naturally-occurring mutations in SARS-CoV-2 variants. N501Y substitution is found in alpha, beta, gamma and omicron variants (Figure 1a) [11]. E484 is mutated in BriLife® to aspartic acid to form E484D. In beta and gamma, E484 is mutated to Lysine (E484K). In omicron, E484 is mutated to Alanine (E484A). E484K, E484D, E484A and E484G substitutions were shown to escape neutralization by convalescent human sera [11], further consolidating the role of E484 in antibody binding. N501Y as a single mutation does not significantly impair sensitivity to vaccine-induced antibodies [12]. However, combination of E484K, N501Y and K417N or K417T found in beta, omicron, or gamma variants, was shown to significantly affect neutralization capacity in sera of convalescent or vaccinated individuals [13]. In omicron, Q493 is mutated to Lysine (Q493K); mutation was previously shown to enable evasion from antibody-mediated immunity [14]. We suggest that the above-mentioned BriLife® mutations might provide an advantage in combating SARS-CoV-2 variants.

P681 located just before the RRAR multibasic motif in furin cleavage site is replaced by Histidine at alpha and omicron variants (P681H) and by Arginine at delta variant (P681R). This region is of major importance to transmissibility and spreading of SARS-CoV-2 [11]. Whether the spontaneously-acquired R685G mutation of BriLife® have a beneficial effect on vaccine efficacy, specifically against alpha, delta, and omicron, awaits further investigation.

As currently approved vaccines correspond to the original virus sequence [4, 15], there is major concern regarding their ability to protect against SARS-CoV-2 variants. Neutralization titers against variants following two-dose vaccination with BNT162b2, mRNA-1273 or ChAdOx1 nCoV-19 were reported, showing for most only a minor reduction in alpha neutralization, but a strong reduction in beta and gamma neutralization [8], while others report no significant reduction in vaccine efficacy against gamma variant [4, 10, 13]. An overall reduction in neutralization potential has been reported against beta variant ranging between ∼2-30-fold, depending on the report [10, 11, 13, 16]. For delta variant, some report a modest reduction in the effectiveness of two-dose vaccines compared to the alpha variant or to the original virus. An overall reduction ranges between ∼2-11 fold [3, 4, 10, 16].

As for omicron, there are increasing reports on its escape from therapeutic monoclonal antibodies, convalescent patients’ antibodies, or vaccine-induced antibodies. The reduction in omicron neutralization by sera post-immunization was reported more so following two-dose vaccination, but also following three vaccine doses, to a lesser extent [9, 17-19]. Several studies addressing neutralization capacity of VOCs in sera obtained at early time points, ranging between 10 days to 3 months following two dose administration of mRNA or adenoviral vaccines show a significant and profound neutralization reduction compared to the original virus. Carreño et al show a 23.3-fold reduction for BNT162b2 at 14-21 days post 2^nd^ vaccination, and a 42.6-fold reduction for mRNA-1273 at 14-36 days post 2^nd^ vaccination [20]; Cele report a ∼22-fold reduction for BNT162b2 at 10-33 days post 2^nd^ vaccination [21]. An additional study shows a ∼108-fold decrease and 18.9-fold decrease between omicron and the original virus at 28 days following 2nd vaccination with BNT162b2, or ADZ1222, respectively [17]. The time points addressed by these reports span the 14 days-time point tested in the current study.

Taken together, most of the above-mentioned studies show a significant reduction following 2^nd^ dose of vaccination against beta, delta, and omicron, already at early time points, which span the 14 days-time point tested in the current study. Our data show only a mild reduction, of 2.4-and 2.9-fold for beta and omicron, respectively. Thus, we suggest that the maintenance of neutralization capacity against variants at these early time point indicates the potential of BriLife® as an efficacious vaccine against variants. It would be interesting to asses neutralization capacity of BriLife® vaccinees’ sera obtained at later time points in future work.

Significant reduction in neutralization potential of convalescent sera from early pandemic stages against variants was also reported [8, 10, 22]. We also compared neutralization capacity of 9 convalescent sera from early pandemic infections (Figure S1) against the original virus and alpha, beta, gamma or delta variants. We show no significant difference in neutralization titers of the original virus and that of alpha, beta or gamma variants (Supplementary Figure S1a-d, Table S4). However, we do see a significant 3.8-fold reduction in neutralization titers of convalescent sera against delta VOC compared to the original virus (Supplementary Figure S1d, Table S4).

Overall, it appears that beta is the most resistant variant to neutralization either by convalescent sera, currently available vaccines, or BriLife® vaccination [8]. Recent cross-neutralization data from convalescent sera from different pandemic waves shed light on the possible contribution of specific spike mutations to protection from variants [8] and gives rise to a need for updated vaccines. Convalescent sera also show lower, or no neutralization of omicron VOC, depending on the variant of infection and on time post infection [9, 19].

Taken together, our data indicates that BriLife®-induced antibodies maintain neutralizing potential against all tested variants, and most importantly against delta, and the recently emerged omicron VOCs. We suggest that spontaneously-acquired mutations that occurred during BriLife® development and correspond to naturally-occurring mutations of SARS-CoV-2 variants, may increase the potential of BriLife® to maintain effectiveness against current SARS-CoV-2 variants, and potentially against future VOC.

## Data Availability

All data produced in the present work are contained in the manuscript

## SUPPLEMENTARY MATERIAL

**Table S1.** Neutralizing titers (NT_50_) of BriLife® vaccinees against SARS-CoV-2 original virus, variants alpha, beta, and gamma (cohort 1). Samples that were below LOD were assigned a value of 10 (half of the LOD).

| **Vaccinee #** | **original virus** | **alpha** | **beta** | **gamma** |
| --- | --- | --- | --- | --- |
| 1 | 241 | 72 | 10 | 321 |
| 2 | 65 | 10 | 44 | 114 |
| 3 | 137 | 54 | 10 | 160 |
| 4 | 197 | 154 | 75 | 228 |
| 5 | 251 | 108 | 34 | 504 |
| 6 | 220 | 49 | 41 | 426 |
| 7 | 240 | 86 | 234 | 349 |
| 8 | 258 | 95 | 60 | 220 |
| 9 | 608 | 836 | 420 | 896 |
| **Mean titer** | **246** | **163** | **103** | **357** |
| Fold change<br>(relative to<br>original virus) |  | 1.5 | 2.4 | 0.7 |

**Table S2.**
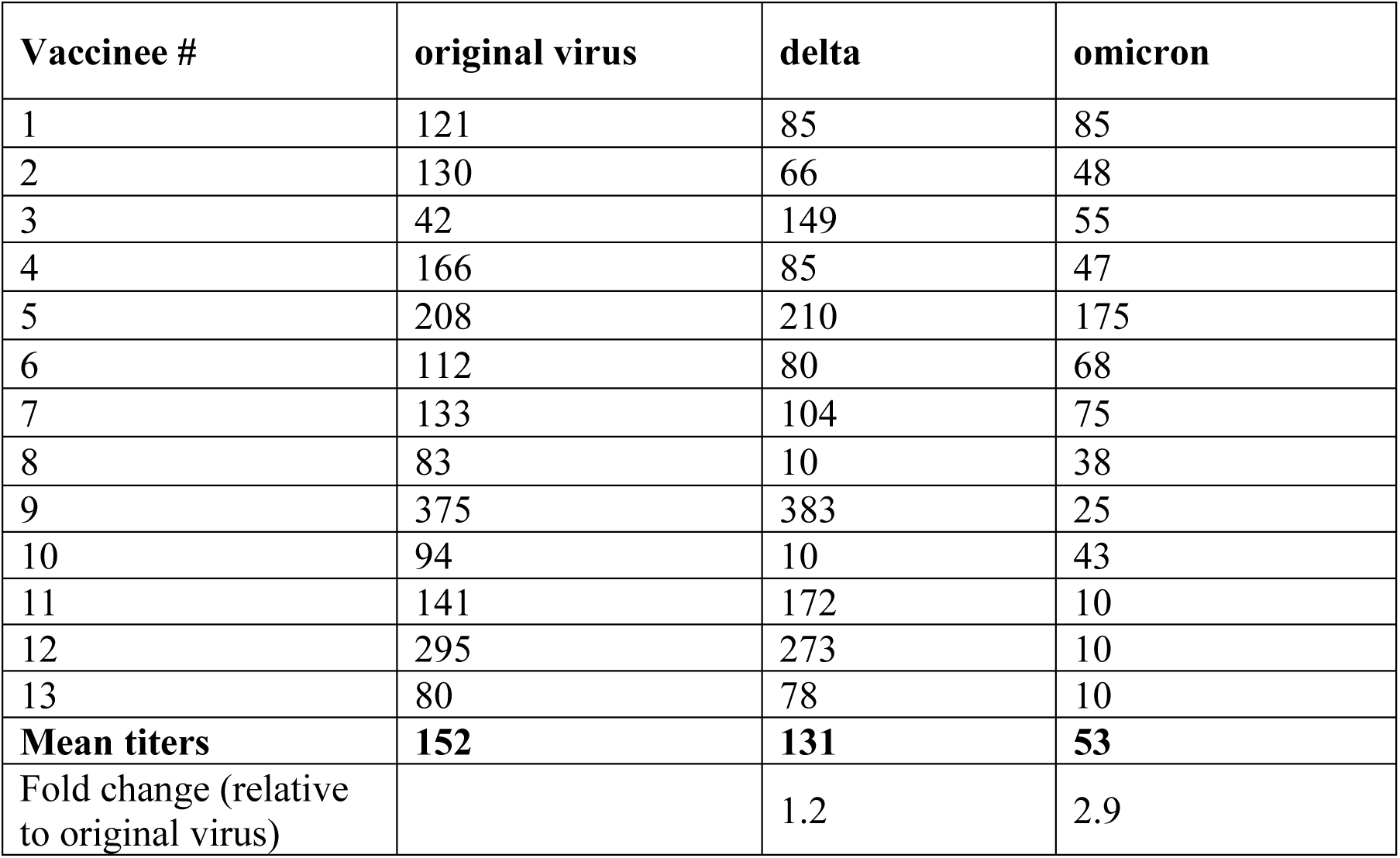
Neutralization titers (NT_50_) of BriLife® vaccinees against SARS-CoV-2 original virus, and delta and omicron variant of concern (cohort 2). Samples that were below LOD were assigned a value of 10 (half of the LOD).

| **Vaccinee #** | **original virus** | **delta** | **omicron** |
| --- | --- | --- | --- |
| 1 | 121 | 85 | 85 |
| 2 | 130 | 66 | 48 |
| 3 | 42 | 149 | 55 |
| 4 | 166 | 85 | 47 |
| 5 | 208 | 210 | 175 |
| 6 | 112 | 80 | 68 |
| 7 | 133 | 104 | 75 |
| 8 | 83 | 10 | 38 |
| 9 | 375 | 383 | 25 |
| 10 | 94 | 10 | 43 |
| 11 | 141 | 172 | 10 |
| 12 | 295 | 273 | 10 |
| 13 | 80 | 78 | 10 |
| **Mean titers** | **152** | **131** | **53** |
| Fold change (relative to original virus) |  | 1.2 | 2.9 |

**Table S3.** Neutralization titers (NT_50_) of BriLife® vaccinees against rVSV-WT (Indiana serotype) at time zero (T0), 28 days post 1^st^ vaccination (P) and two weeks following 2^nd^ vaccination (P+B). Samples that were below LOD were assigned a value of 10 (half of the LOD).

| **Vaccinee #** | **T0** | **P** | **P+B** |
| --- | --- | --- | --- |
| 1 | 21 | 38 | 62 |
| 2 | 83 | 99 | 202 |
| 3 | 10 | 10 | 72 |
| 4 | 10 | 10 | 10 |
| 5 | 115 | 57 | 52 |
| 6 | 10 | 45 | 10 |
| 7 | 10 | 10 | 10 |
| 8 | 43 | 30 | 45 |
| 9 | 10 | 10 | 59 |
| 10 | 10 | 10 | 10 |
| **Mean titers** | **32** | **32** | **53** |

**Table S4.** Neutralizing titers (NT_50_) of convalescent sera against SARS-CoV-2 original virus, variants alpha, beta, gamma and delta (cohort 3). Samples that were below LOD were assigned a value of half of the LOD.

| **Convalescent sera #** | **original virus** | **alpha** | **beta** | **gamma** | **delta** |
| --- | --- | --- | --- | --- | --- |
| 1 | 541 | 296 | 279 | 423 | 85 |
| 2 | 1385 | 1027 | 760 | 953 | 212 |
| 3 | 758 | 197 | 139 | 333 | 160 |
| 4 | 142 | 177 | 160 | 95 | 79 |
| 5 | 366 | 100 | 107 | 297 | 178 |
| 6 | 323 | 40 | 79 | 269 | 115 |
| 7 | 119 | 41 | 65 | 132 | 39 |
| 8 | 104 | 20 | 66 | 40 | 103 |
| 9 | 123 | 51 | 39 | 50 | 41 |
| **Mean titers** | **429** | **216** | **188** | **288** | **113** |
| Fold change (relative to original virus) |  | 2.0 | 2.3 | 1.5 | 3.8 |

**Figure S1:**
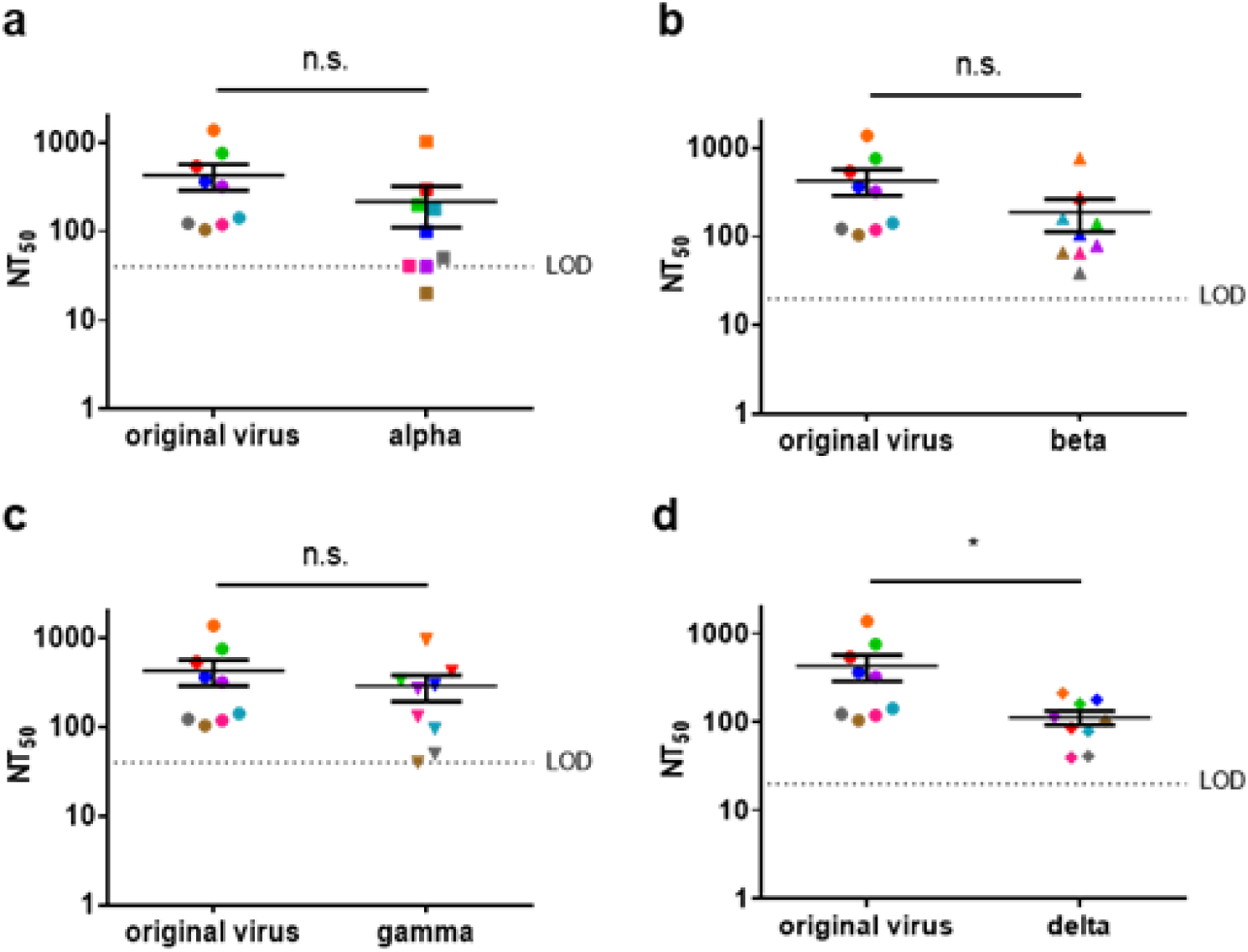
Neutralization of alpha, beta, gamma and delta variants by sera from convalescent patients: (a) NT_50_ of convalescent patients’ sera against SARS-CoV-2 original virus, in comparison to (a) alpha, (b) beta, (c) gamma and (d) delta (n=9) variants. Each serum is color coded for all tested viruses. LOD is indicated in each panel (a,c LOD=40; b,d LOD=20). Samples below LOD were assigned a value of half the LOD. Data is presented as mean ± SEM. Significance was determined by unpaired t-test. p values: 0.2454, 0.1514, 0.4184, 0.0409, for comparison of original virus to alpha, beta, gamma and delta, respectively.

## Author Contributions

Conceptualization, Y.Y.R., N.E., T.I., N.P. ; methodology, Y.Y.R., N.E., T.I., N.P. ; formal analysis, Y.Y.R., N.E. ; investigation, Y.Y.R., N.E., M.F., H.T., B.P., H.A., S.M., I.G., A.M., S.W., I.C.G., O.I., A.B.D., A.Z. ; resources, M.M., M.I., D.C., E.S., H.M., N.M.B., Y.C., R.Z. ; writing—original draft preparation, Y.Y.R., T.I., N.P. ; writing—review and editing, Y.Y.R, T.I., N.P. ; supervision, T.I., N.P. ; All authors have read and agreed to the published version of the manuscript.

## Funding

This research received no external funding.

## Institutional Review Board Statement

BriLife® study - was performed in compliance with International Council for Harmonization (ICH) Good Clinical Practices (GCP), including the archiving of essential documents as well as the ethical principles of the Declaration of Helsinki. Convalescent COVID-19 sera - The study was approved by the ethics committee of the Israeli Ministry of Health (0083-20-WOMC)^2^.

## Informed Consent Statement

Informed consent was obtained from all BriLife® participants involved in the study. All convalescent volunteers gave their informed consent to the National Blood services of Magen David Adom.

## Data Availability Statement

Data available on request due to restrictions, e.g., privacy or ethical.

## Acknowledgments

We thank the participants in BriLife® clinical trial for their altruism and their dedication to this trial. We thank Prof. Yoseph Caraco from Hadassah Medical Center (Jerusalem), Dr. Eytan Ben-Ami from Sheba Medical Center (Ramat Gan), Dr. Avivit Peer from Rambam Health Care Campus (Haifa), Prof. David Zeltser from Tel Aviv Sourasky Medical Center (Tel Aviv), Prof. Shlomo Maayan from Barzilai Medical Center (Ashkelon), Dr. Victor Vishlitzky from Meir Medical Center (Kefar Sava), Dr. Tal Brosh from Assuta (Ashdod), Dr. Noa Eliakim from Rabin Medical Center (Petah Tiqwa), and the respective Clinical Center teams in Israel participating in this study. We also acknowledge the contributions of the IIBR20-001 Clinical Trial Group at IIBR, Gsap (Haifa, Israel) and IQVIA (Durham NC, USA).

SARS-CoV-2 original virus was kindly provided by Bundeswehr Institute of Microbiology, Munich, Germany. SARS-CoV-2 variants alpha, beta, gamma and delta were kindly provided by Prof. Ella Mendelson, Dr. Ital Nemet and Limor Kliker from the Central Virology Lab of the Israel Ministry of Health. rVSV-WT was kindly provided by Prof. Eran Bacharach from Tel Aviv University.

## Conflicts of Interest

The authors declare no conflict of interest.

## Notes

### Competing Interest Statement

The authors have declared no competing interest.

### Clinical Trial

NCT04608305

### Funding Statement

This study did not receive any funding

### Author Declarations

Serum samples of BriLife® vaccinees were obtained from participants in a randomized, multi-center, placebo-controlled, dose-escalation phase II of an ongoing clinical trial, aimed to evaluate the safety, immunogenicity and potential efficacy of BriLife®, an rVSV-SARS-CoV-2-S vaccine (IIBR-100) in Adults (ClinicalTrials.gov - NCT04608305). This study was performed in compliance with International Council for Harmonisation (ICH) Good Clinical Practices (GCP), including the archiving of essential documents as well as the ethical principles of the Declaration of Helsinki. Serum samples of convalescent COVID-19 patients were collected by the National Blood Services of Magen David Adom in Israel within a protocol for plasma donation. All convalescent volunteers gave their informed consent to the National Blood services of Magen David Adom. The study was approved by the ethics committee of the Israeli Ministry of Health (0083-20-WOMC).

### Summary of Updates

Omicron VOC data was added

## References

1. Yahalom-Ronen, Y., et al., A single dose of recombinant VSV-G-spike vaccine provides protection against SARS-CoV-2 challenge. Nat Commun, 2020. 11(1): p. 6402.

2. Maor, Y., et al., Compassionate use of convalescent plasma for treatment of moderate and severe pneumonia in COVID-19 patients and association with IgG antibody levels in donated plasma. EClinicalMedicine, 2020. 26: p. 100525.

3. Lustig, Y., et al., Neutralising capacity against Delta (B.1.617.2) and other variants of concern following Comirnaty (BNT162b2, BioNTech/Pfizer) vaccination in health care workers, Israel. Euro Surveill, 2021. 26(26).

4. Davis, C., et al., Reduced neutralisation of the Delta (B.1.617.2) SARS-CoV-2 variant of concern following vaccination. PLoS Pathog, 2021. 17(12): p. e1010022.

5. Levine-Tiefenbrun, M., et al., Viral loads of Delta-variant SARS-CoV2 breakthrough infections following vaccination and booster with the BNT162b2 vaccine. medRxiv, 2021: p. 2021.08.29.21262798.

6. Kuhlmann C M.C., Claassen M, Maponga T, Burgers WA, Keeton R, Riou C, Sutherland AD, Suliman T, Shaw ML, Preiser W., Breakthrough infections with SARS-CoV-2 omicron despite mRNA vaccine booster dose. Lancet., 2022.

7. Marzi, A., et al., Vesicular stomatitis virus-based vaccines against Lassa and Ebola viruses. Emerg Infect Dis, 2015. 21(2): p. 305–7.

8. Liu, C., et al., Reduced neutralization of SARS-CoV-2 B.1.617 by vaccine and convalescent serum. Cell, 2021. 184(16): p. 4220–4236 e13.

9. Planas, D., et al., Considerable escape of SARS-CoV-2 Omicron to antibody neutralization. Nature, 2021.

10. Planas, D., et al., Reduced sensitivity of SARS-CoV-2 variant Delta to antibody neutralization. Nature, 2021. 596(7871): p. 276–280.

11. Harvey, W.T., et al., SARS-CoV-2 variants, spike mutations and immune escape. Nat Rev Microbiol, 2021. 19(7): p. 409–424.

12. Pegu, A., et al., Durability of mRNA-1273 vaccine-induced antibodies against SARS-CoV-2 variants. Science, 2021. 373(6561): p. 1372–1377.

13. Hoffmann, M., et al., SARS-CoV-2 variants B.1.351 and P.1 escape from neutralizing antibodies. Cell, 2021. 184(9): p. 2384–2393 e12.

14. Thomson, E.C., et al., Circulating SARS-CoV-2 spike N439K variants maintain fitness while evading antibody-mediated immunity. Cell, 2021. 184(5): p. 1171–1187 e20.

15. Puranik, A., et al., Comparison of two highly-effective mRNA vaccines for COVID-19 during periods of Alpha and Delta variant prevalence. medRxiv, 2021.

16. Noori, M., et al., Potency of BNT162b2 and mRNA-1273 vaccine-induced neutralizing antibodies against severe acute respiratory syndrome-CoV-2 variants of concern: A systematic review of in vitro studies. Rev Med Virol, 2021: p. e2277.

17. Dejnirattisai, W., Huo, J., Zhou, D., Zahradník, J.?í, Supasa, P., Liu, C.,, et al., SARS-CoV-2 Omicron-B.1.1.529 leads to widespread escape from neutralizing antibody responses. Cell, 2022.

18. Nemet, I., et al., Third BNT162b2 Vaccination Neutralization of SARS-CoV-2 Omicron Infection. N Engl J Med, 2021.

19. Ikemura, N., et al., SARS-CoV-2 Omicron variant escapes neutralization by vaccinated and convalescent sera and therapeutic monoclonal antibodies. medRxiv, 2021.

20. Carreno, J.M., et al., Activity of convalescent and vaccine serum against SARS-CoV-2 Omicron. Nature, 2021.

21. Cele, S., et al., Omicron extensively but incompletely escapes Pfizer BNT162b2 neutralization. Nature, 2021.

22. Wang, B., et al., Resistance of SARS-CoV-2 variants to neutralization by convalescent plasma from early COVID-19 outbreak in Singapore. NPJ Vaccines, 2021. 6(1): p. 125.

